# Effectiveness of COVID-19 vaccines against the Omicron (B.1.1.529) variant of concern

**DOI:** 10.1101/2021.12.14.21267615

**Authors:** Nick Andrews, Julia Stowe, Freja Kirsebom, Samuel Toffa, Tim Rickeard, Eileen Gallagher, Charlotte Gower, Meaghan Kall, Natalie Groves, Anne-Marie O’Connell, David Simons, Paula B. Blomquist, Asad Zaidi, Sophie Nash, Nurin Iwani Binti Abdul Aziz, Simon Thelwall, Gavin Dabrera, Richard Myers, Gayatri Amirthalingam, Saheer Gharbia, Jeffrey C. Barrett, Richard Elson, Shamez N Ladhani, Neil Ferguson, Maria Zambon, Colin NJ Campbell, Kevin Brown, Susan Hopkins, Meera Chand, Mary Ramsay, Jamie Lopez Bernal

## Abstract

**Background:** A rapid increase in cases due to the SARS-CoV-2 Omicron (B.1.1.529) variant in highly vaccinated populations has raised concerns about the effectiveness of current vaccines.

**Methods:** We used a test-negative case-control design to estimate vaccine effectiveness (VE) against symptomatic disease caused by the Omicron and Delta variants in England. VE was calculated after primary immunisation with two BNT162b2 or ChAdOx1 doses, and at 2+ weeks following a BNT162b2 booster.

**Results:** Between 27 November and 06 December 2021, 581 and 56,439 eligible Omicron and Delta cases respectively were identified. There were 130,867 eligible test-negative controls. There was no effect against Omicron from 15 weeks after two ChAdOx1 doses, while VE after two BNT162b2 doses was 88.0% (95%CI: 65.9 to 95.8%) 2-9 weeks after dose 2, dropping to between 34 and 37% from 15 weeks post dose 2.From two weeks after a BNT162b2 booster, VE increased to 71.4% (95%CI: 41.8 to 86.0%) for ChAdOx1 primary course recipients and 75.5% (95%CI: 56.1 to 86.3%) for BNT162b2 primary course recipients.

For cases with Delta, VE was 41.8% (95%CI: 39.4-44.1%) at 25+ weeks after two ChAdOx1 doses, increasing to 93.8% (95%CI: 93.2-94.3%) after a BNT162b2 booster. With a BNT162b2 primary course, VE was 63.5% (95%CI: 61.4 to 65.5%) 25+ weeks after dose 2, increasing to 92.6% (95%CI: 92.0-93.1%) two weeks after the booster.

**Conclusions:** Primary immunisation with two BNT162b2 or ChAdOx1 doses provided no or limited protection against symptomatic disease with the Omicron variant. Boosting with BNT162b2 following either primary course significantly increased protection.

## Introduction

On 26 November 2021 the World Health Organization Technical Advisory Group on SARS-CoV-2 Virus Evolution named the B.1.1.529 COVID-19 variant, first detected in Botswana and South Africa, as the Omicron variant of concern.(1) This classification was based on a rapid increase in confirmed SARS-CoV-2 infection in South Africa, coinciding with an increase in detections of the Omicron variant, identification of a number of concerning mutations and early evidence of an increased risk of reinfection in recently-infected individuals.

A large number of mutations have been identified in the Omicron variant, including multiple mutations in the receptor binding domain of the spike protein which have been associated with increased transmissibility and immune evasion after natural infection and vaccination.(2) Emerging laboratory data indicate significantly reduced neutralising antibody response to Omicron compared to the original COVID-19 virus or the Delta variant in vaccinated individuals, although booster doses improved neutralising activity.(3, 4) Neutralising antibody has been found to correlate with protection against reinfection and vaccine effectiveness against infection, therefore reduced vaccine effectiveness against Omicron is anticipated based on these early laboratory findings.(5-7)

COVID-19 vaccines are highly effective against symptomatic disease and, more so, against severe disease and fatal outcomes with the original strain as well as the Alpha variant that predominated in early 2021.(8-14) Modest reductions in vaccine effectiveness against infection and mild disease have been observed with Beta and Delta variants, although effectiveness against severe disease has remained high for at least 6 months after primary immunisation with two COVID-19 vaccine doses.(15-18) Waning of protection has been observed with time since vaccination, and especially with the Delta variant which is able to at least partially evade natural and vaccine-induced immunity.(19) However, third (booster) doses provide a rapid and significant increase in protection against both mild and severe disease outcomes.(18, 20-24)

In the UK, Omicron cases were first identified in mid-November 2021 through whole genome sequencing of polymerase chain reaction (PCR) positive swabs. Initially cases occurred primarily in travellers and their close contacts but there was already evidence of community transmission from late November.(25) The UK COVID-19 vaccination programme has been in place since December 2020 with primary courses of two doses of either BNT162b2 (Pfizer-BioNTech, Comirnaty®), ChAdOx1-S (Vaxzevria, AstraZeneca) or mRNA-1273 (Spikevax, Moderna). Two dose vaccine uptake is more than 60% in all cohorts over 20 years and over 80% in all cohorts over 50 years of age, with vaccinations now being offered to children over the age of 12 years.(26) Booster vaccination with either BNT162b2 or a half dose (50µg) of mRNA-1273 was introduced in September 2021 to adults over 50 years and those in risk groups, and later expanded to all adults. Initially boosters were offered 6 months after completion of the primary course. With the emergence of the Omicron variant, this interval was reduced to 3 months.(27)

In this study we estimate vaccine effectiveness against symptomatic disease after 2 doses of BNT162b2 and ChAdOx1-S, and after booster doses of BNT162b2 following a primary immunisation with either BNT162b2 or ChAdOx1-S.

## Methods

### Study Design

A test negative case control design was used to estimate vaccine effectiveness against symptomatic COVID-19 with the Omicron variant compared to the Delta variant.(16) The odds of vaccination in symptomatic PCR positive cases was compared to the odds of vaccination in symptomatic individuals who tested negative for SARS-CoV-2 in England.

### Data Sources

#### COVID-19 Testing Data

SARS-CoV-2 Testing PCR testing for SARS CoV-2 in England is undertaken by hospital and public health laboratories (Pillar 1), as well as by community testing (Pillar 2) including drive through and at-home testing, which is available to anyone with symptoms consistent with COVID-19 (high temperature, new continuous cough, or loss or change in sense of smell or taste), is a contact of a confirmed case, for care home staff and residents or who have self-tested as positive using a lateral flow device. SARS-CoV-2 lateral flow tests (LFT) are freely available to all members of the population for regular home testing. Data on all positive PCR and LFT tests, and on negative Pillar 2 PCR tests from symptomatic individuals with an onset date after 16th October 2020 were extracted up to 6th December 2021. Individuals who reported symptoms and were tested in Pillar 2 between November 27^th^ and December 3^rd^ 2021 were included in the analysis. Any negative tests taken within 7 days of a previous negative test, or where symptoms were recorded, with symptoms within 10 days of symptoms for a previous negative test were dropped as these likely represent the same episode. Negative tests taken within 21 days before a positive test were also excluded as there is a high chance of these being false negatives. Positive and negative tests within 90 days of a previous positive test were also excluded, however where participants had later positive tests within seven days of a positive then preference was given to PCR tests, symptomatic tests and tests with sequencing or S-gene testing done. Data were restricted to persons who had reported symptoms and gave an onset date within the 10 days prior to testing to account for reduced PCR sensitivity beyond this period in an infection event. Data from cases who had reported recent travel were excluded due to differences in exposure risk and possible misclassification of vaccination status. Only samples tested in one of the community testing laboratories using the TaqPath assay were included in the final analysis, and only positive cases with S-gene test or sequencing results available were included. A small number of positive tests were excluded where sequencing found them to be neither the Delta nor Omicron variant. Finally, only samples taken from November 27th were retained for analysis as this corresponded to the period when S-gene negative status was predictive of being the Omicron variant.

#### Vaccination Data

The National Immunisation Management System (NIMS) contains demographic information on the whole population of England who are registered with a GP in England and is used to record all COVID-19 vaccinations.(28) NIMS was accessed on 09 December 2021 for dates of vaccination and manufacturer, sex, date of birth, ethnicity, and residential address.(29) Addresses were used to determine index of multiple deprivation quintile and were also linked to Care Quality Commission registered care homes using the unique property reference number. Data on geography (NHS region), risk group status, clinically extremely vulnerable status, and health/social care worker were also extracted from the NIMS. Booster doses were identified as a third dose given at least 140 days after a second dose and administered after 13 September 2021. Individuals with four or more doses of vaccine, heterologous primary schedule or fewer than 19 days between their first and second dose were excluded.

#### Identification of Delta and Omicron Variants and assignment to cases

Sequencing of PCR positive samples is undertaken through a network of laboratories, including the Wellcome Sanger Institute. Whole-genome sequences are assigned to UKHSA definitions of variants based on mutations. Spike gene (S-gene) target status on PCR-testing is a second approach for identifying each variant because the Omicron variant has been associated with a negative S-gene target (S-gene target failure, SGTF) on PCR testing with the Taqpath assay whereas the Delta variant almost always has a positive S-gene target.(25) Approximately 40% of Pillar 2 Community testing in England is carried out by laboratories using the TaqPath assay (Thermo Fisher Scientific). Cases were defined as the Delta or Omicron variant based on whole genome sequencing or S-gene target status, with sequencing taking priority. A priori, we considered that S-gene target failure (SGTF) would be used to define the Omicron variant when Omicron accounts for at least 80% of S-gene target failure cases.

Testing data were linked to NIMS on 09 December 2021 using combinations of the unique individual National Health Service (NHS) number, date of birth, surname, first name, and postcode using deterministic linkage - 92.2% of eligible tests could be linked to the NIMS.

### Statistical Analysis

Logistic regression was used, with the PCR test result as the dependent variable and cases being those testing positive (stratified in separate analyses as either Omicron or Delta) and controls being those testing negative. Vaccination status was included as an independent variable and effectiveness defined as 1-odds of vaccination in cases/odds of vaccination in controls.

Vaccine effectiveness was adjusted in logistic regression models for age (5 year bands up to age 60, then everyone age 60+), sex, index of multiple deprivation (quintile), ethnic group, geographic region (NHS region), period (day of test), health and social care worker status, clinical risk group status, clinically extremely vulnerable, and previously testing positive. These factors were all considered potential confounders so were included in all models.

Analyses were stratified by primary immunisation course (ChAdOx1-S or BNT162b2). Any heterologous primary schedules were excluded. Vaccine effectiveness was assessed for each primary course in intervals of 2-9, 10-14, 15-19, 20-24 and 25+ weeks post dose 2. Vaccine effectiveness was also assessed at least two weeks after a BNT162b2 booster for primary immunisation with ChAdOx1-S or BNT162b2. Comparison was to unvaccinated individuals to estimate the absolute effectiveness of vaccination against Omicron and Delta variants.

Numbers were too small to estimate vaccine effectiveness with mRNA-1273 as either a primary course or a booster.

## Results

Within sequenced cases from Pillar 2 testing where S-gene testing was done the proportion of S-gene negative tests that were sequenced as Omicron was 6/12 (50%) on 25^th^ November, 13/20 (65%) on 26^th^ November, 18/20 (90%) on 27^th^ November, 10/11 (91%) on 28^th^ November and 17/19 (89%) on 29^th^ November. We therefore used cases tested from 27^th^ November where the positive predictive value was above 80%. Sequencing or S-gene target status were both used to identify Omicron and Delta cases, with the sequencing result taking priority.

There were 581 symptomatic Omicron cases which were identified during the study period by sequencing or SGTF and linked to NIMS for vaccination status. Over the same period there were 56,439 eligible Delta cases and 130,867 test negative controls. A description of the eligible tests is given in Table 1. Omicron cases 14+ days after a booster occurred a median of 41 days after the booster (range 14-72 days).

**Table 1:**
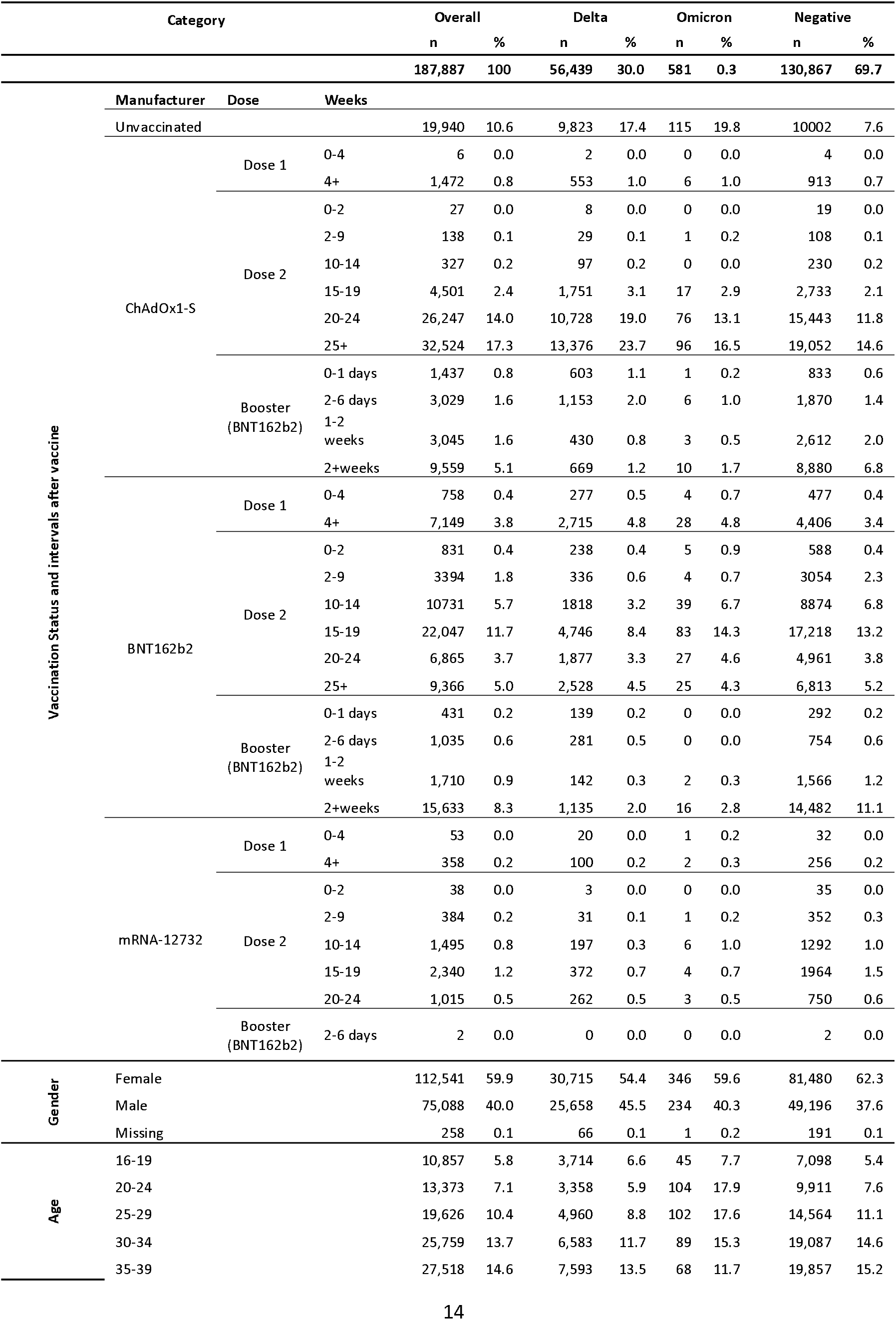
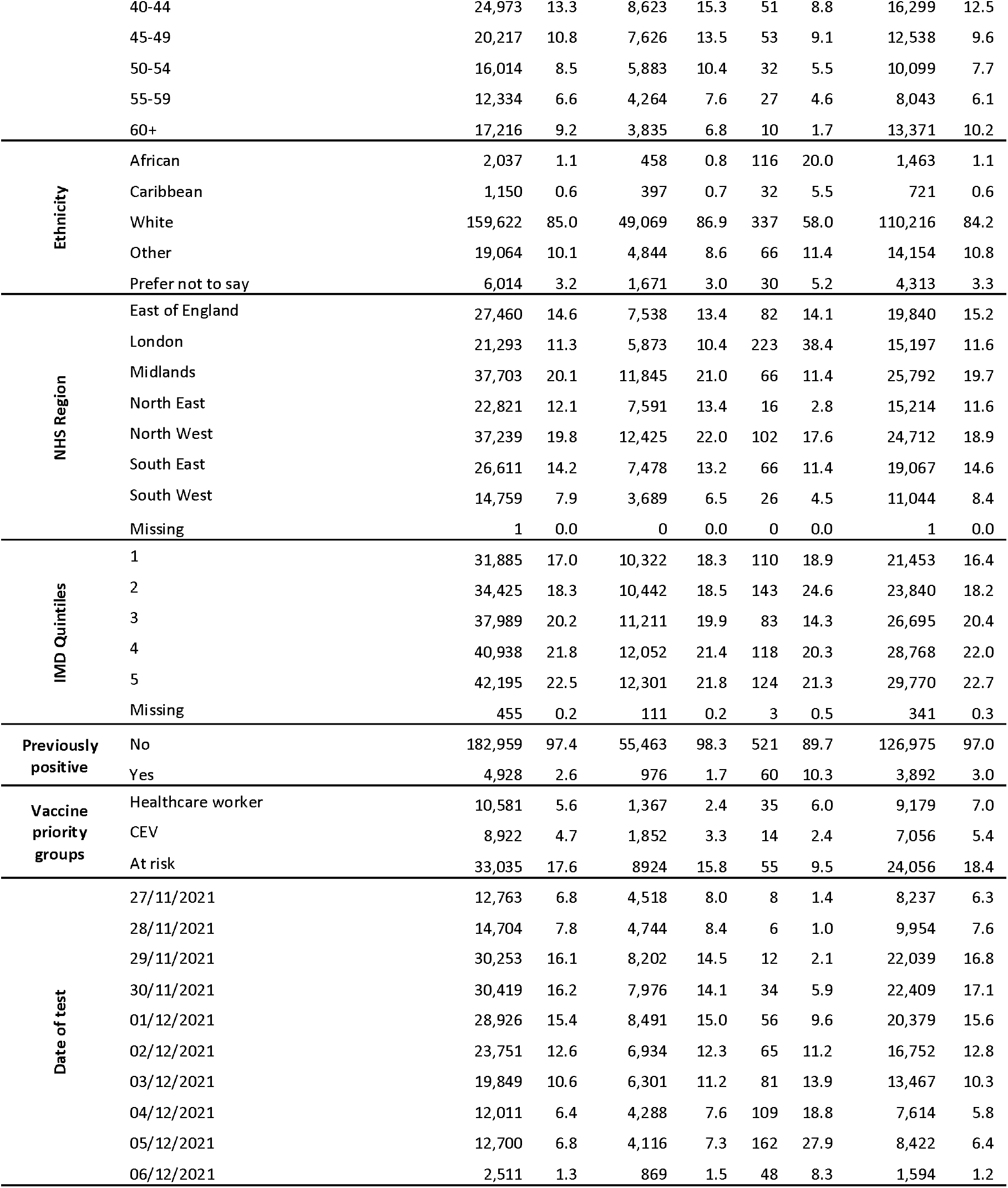
Descriptive characteristics of positive and negative test results in individuals tested for SARS-CoV-2 in England for the study population

Vaccine effectiveness against symptomatic disease in individuals who received a primary course of the ChAdOx1 or BNT162b2 is shown by period after primary immunisation with 2 doses and after a third dose with BNT162b2 is shown in Figure 1 and Table 2. Apart from 2-9 weeks post dose 2 for BNT162b2, effectiveness was lower for Omicron compared to Delta post vaccination at all time interval investigated. Among those who had received two ChAdOx1 doses, there was no protective effect of vaccination against symptomatic disease with Omicron from 15 weeks after the second dose. Among those who had received two BNT162b2 doses, vaccine effectiveness was 88.0% (95%CI: 65.9 to 95.8%) 2-9 weeks after dose 2, dropping to 48.5% (95%CI: 24.3 to 65.0%) at 10-14 weeks post dose 2 and dropping further to between 34 and 37% from 15 weeks post dose 2. Among those who received ChAdOx1 as the primary course, from 2 weeks after a BNT162b2 booster dose, vaccine effectiveness increased to 71.4% (95%CI: 41.8 to 86.0%). Vaccine effectiveness increased to 75.5% (95%CI: 56.1 to 86.3%) after the booster among those who had received BNT162b2 as the primary course.

**Table 2:**
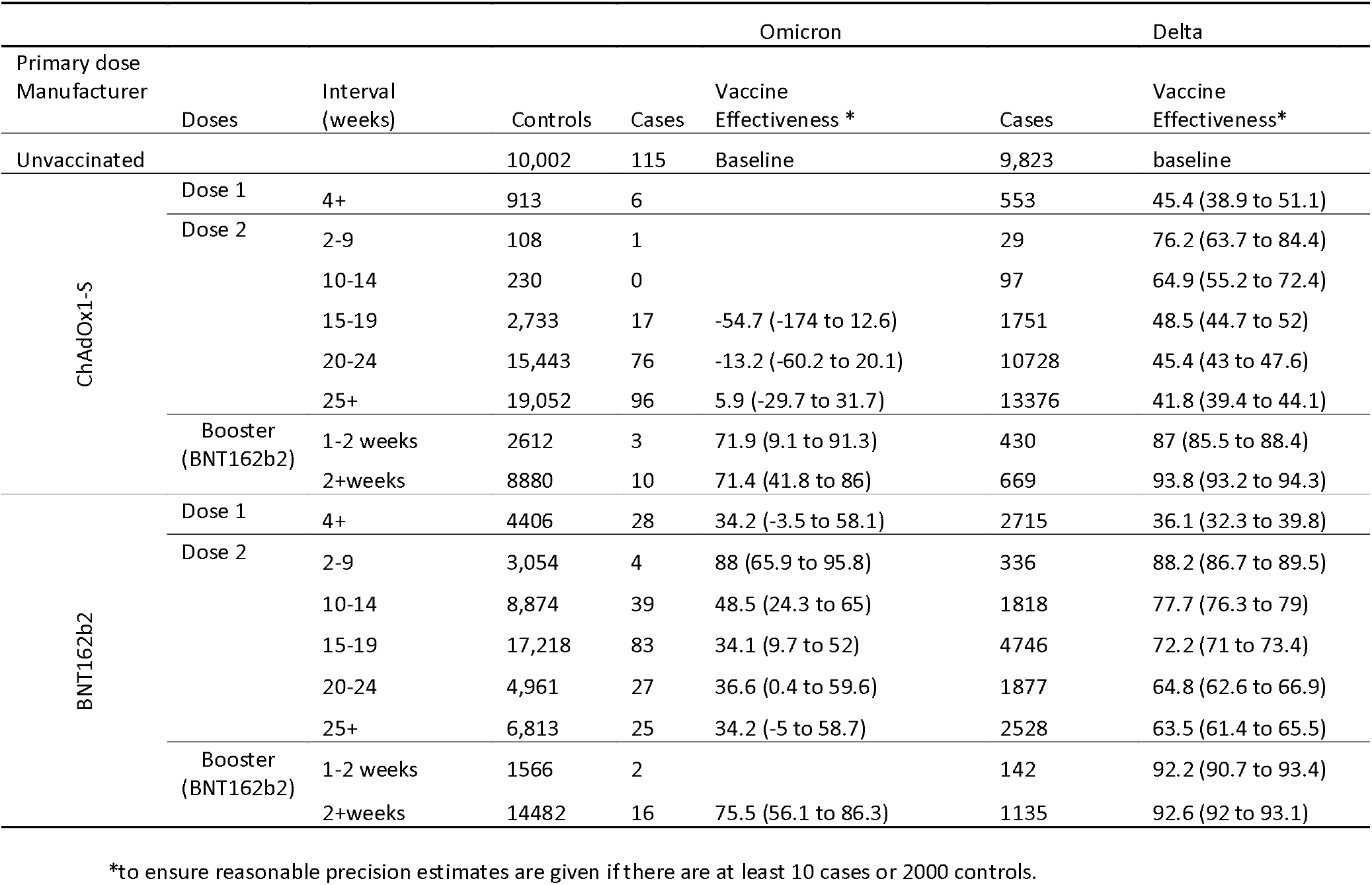
Vaccine effectiveness (%) against symptomatic diseases by period after dose 1 and dose 2 for Delta and Omicron*

**Figure 1:**
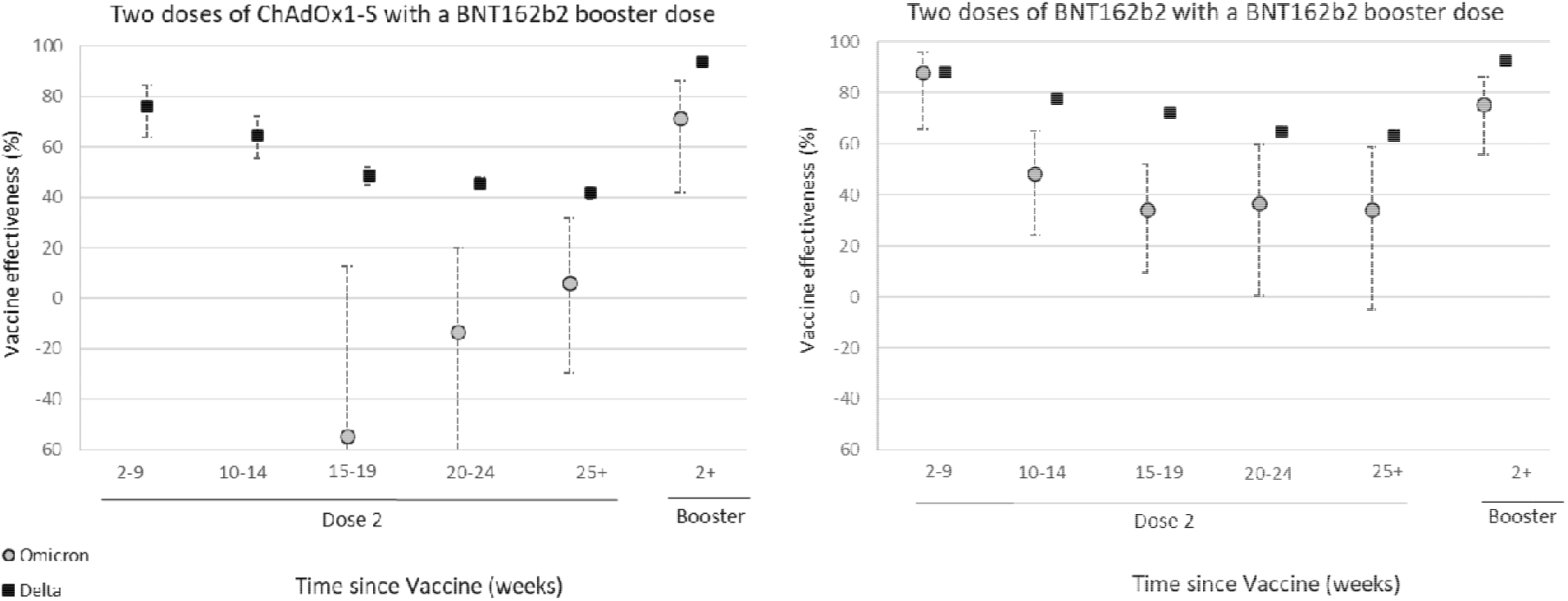
Vaccine effectiveness against symptomatic diseases by period after dose 2 and a booster dose for Delta (black squares) and Omicron (grey circles) for recipients of two doses of ChAdOx1-S as the primary course and BNT162b2 as a booster and for recipients of two doses of BNT162b2 as the primary course and BNT162b2as a booster

With the Delta variant, effectiveness dropped from 76.2% (95%CI: 63.7 to 84.4%) 2-9 weeks after dose 2 down to 41.8% (95%CI: 39.4-44.1%) at 25+ weeks after dose 2 with a ChAdOx1 primary course. Effectiveness increases to 93.8% (95%CI: 93.2-94.3%) two weeks after a BNT162b2 booster. With a BNT162b2 primary course, effectiveness drops from 88.2% (95%CI: 86.7 to 89.5%) 2-9 weeks after dose 2 down to 63.5% (95%CI: 61.4 to 65.5%) 25+ weeks after dose 2, increasing to 92.6% (95%CI: 92.0-93.1%) two weeks after the booster.

All vaccine effectiveness estimates for the Omicron variant have with wide confidence intervals because of small case numbers.

## Discussion

Our findings indicate that vaccine effectiveness against symptomatic disease with the Omicron variant is significantly lower than with the Delta variant. We are unable to determine protection against severe forms of disease due to the small number of Omicron cases so far and the natural lag between infection and more severe outcomes. Previous UK experience with the Delta variant suggested that protection against hospitalisation after two doses of vaccine was relatively well maintained.(18) Despite the low effectiveness in the longer intervals after primary vaccination shown here, moderate to high vaccine effectiveness against mild infection of 70-75% was seen in the early period after a booster dose of BNT162b2 following either ChAdOx1-S or BNT162b2 as a primary course.

These findings are consistent with preliminary neutralisation data for the Omicron variant. The South African and German studies, as well as unpublished UK data indicate a 20 to 40 fold reduction in neutralising activity in sera from two dose BNT162b2 vaccine recipients compared to early pandemic viruses and at least a 10 fold reduction compared to the Delta variant.(3, 4, 30) In sera from recipients of two doses of ChAdOx1, a greater reduction in neutralising activity was observed with a high proportion of post-vaccination sera exhibiting neutralising activity below the limit of quantification in the assay.(30) Reassuringly though, higher neutralising activity was observed after a booster dose.(3, 4, 30)

Whilst a correlation between neutralising antibody levels and vaccine effectiveness against symptomatic SARS-CoV-2 infection has been observed at a population level,(5) a similar correlation with effectiveness against severe disease is much less certain. With previous variants, vaccine effectiveness against severe disease, including hospitalisation and death, has been higher and retained for a longer period than effectiveness against mild disease.(15, 18) Cellular immune responses are likely to play a relatively more important role in protection as antibodies wane with time since infection or vaccination; antibody waning may increase the risk of SARS-CoV-2 infection while cellular immunity likely limits progression to severe disease.(31) Cellular immunity is also likely to play a more important role in protection against SARS-CoV-2 variants than antibodies.(31) REF It will be some time before effectiveness against severe disease with Omicron can be estimated but, based on experience with other variants,(18) this is likely to be substantially higher than the estimates against symptomatic disease.

It is important to note that there are differences in the populations that have received different vaccines as a primary course. For example, ChAdOx1 was the main vaccine used early in the programme in care homes and among those in clinical risk groups. Furthermore, mRNA vaccines were the main vaccines used in under 40 year olds following the association between ChAdOx1 and vaccine induced thrombotic thrombocytopenia.(32) While adjustments were made for age and clinical risk factors, this may explain some of the differences in the findings for the primary course – for example the high vaccine effectiveness against Omicron 2-9 weeks after the second dose of BNT162b2 is likely to be primarily among recently vaccinated young adults and teenagers. The early observations for two doses of AstraZeneca are particularly likely to be unreliable as they are based on relatively small numbers and are likely to reflect an older population and a population with more co-morbidities than those given the Pfizer vaccine - this may explain the negative point estimates. There will also be differences in populations that have received a booster dose compared to those who have only received two doses, with the former skewed towards older populations with more comorbidities. Those that have not yet received a booster could have done for reasons that may be associated with exposure risk, for example, booster vaccination may have been delayed due to an outbreak in a closed setting.

The large scale of testing and sequencing in the UK, as well as the use of a national vaccination register has enabled rapid evaluation of vaccine effectiveness against symptomatic infection with the Omicron variant. Nevertheless, there are some limitations and findings should be interpreted with caution. During this early period of circulation of the new variant, a large proportion of cases occurred among travellers. Individuals who reported travel in the preceding two weeks were excluded from this analysis, however, this may not exclude all travellers and will not exclude contacts of travellers. This group is likely to have different exposure to the wider population and may also have different levels of vaccine coverage, therefore residual confounding may be present. Due to the relatively small number of cases of Omicron in the UK, there is significant uncertainty to our estimates and we are unable to break down estimates by population characteristics by that may affect vaccine effectiveness (such as age and clinical risk group).(18) In this analysis, our comparator group is unvaccinated individuals, who comprise a very small proportion of individuals in several age cohorts. These people are likely to differ from the general population according to characteristics that could confound our vaccine effectiveness estimates. In this analysis that covers all ages, this may be less of an issue than in analyses restricted to elderly populations. Furthermore, recent analyses using different control groups have found good concordance when using an unvaccinated control compared to relative vaccine effectiveness between those who have received a booster dose and those who have received two doses. Booster doses have only recently been rolled out in England, therefore we are only able to estimate vaccine effectiveness for a short period after booster vaccination and we do not have information on the duration of protection following a booster dose. There may also be some misclassification due to both imperfect sensitivity and specificity of PCR testing, as well as the use of SGTF to identify Omicron cases.

Our findings indicate that two doses of vaccination with BNT162b2 or ChAdOx1 are insufficient to give adequate levels of protection against infection and mild disease with the Omicron variant, although we cannot comment on protection against severe disease. Boosting with BNT162b2, however, provides a significant increase in protection against mild disease and will likely offer even greater levels of protection against severe and fatal disease. Our findings support maximising coverage with third doses of vaccine in highly vaccinated populations such as the UK. Further follow-up will be needed to assess protection against severe disease and the duration of protection after booster vaccination.

## Data Availability

Data cannot be made publicly available
for ethical and legal reasons, i.e. public availability would compromise patient
confidentiality as data tables list single counts of individuals rather than aggregated

## Acknowledgements

We thank the UK Health Security Agency (UKHSA) covid-19 Data Science Team, UKHSA Outbreak Surveillance Team, NHS England, NHS Digital, and NHS Test and Trace for their roles in developing and managing the COVID-19 testing, variant identification and vaccination systems and datasets as well as reporting NHS vaccinators, NHS laboratories, UKHSA laboratories, and lighthouse laboratories; and we thank the Wellcome Sanger Institute and other laboratories involved in whole genome sequencing of COVID-19 samples; and we thank the Joint Committee on Vaccination and Immunisation and the UK Variant Technical Group for advice and feedback in developing this study.

## Ethics

Surveillance of coronavirus disease 2019 (Covid-19) testing and vaccination is undertaken under Regulation 3 of the Health Service (Control of Patient Information) Regulations 2002 to collect confidential patient information (www.legislation.gov.uk/uksi/2002/1438/regulation/3/made.opensinnewtab) under Sections 3(i) (a) to (c), 3(i)(d) (i) and (ii), and 3. The study protocol was subject to an internal review by the Public Health England Research Ethics and Governance Group and was found to be fully compliant with all regulatory requirements. Given that no regulatory issues were identified and that ethics review is not a requirement for this type of work, it was decided that a full ethics review would not be necessary.

